# Benefit-finding and self-compassion promote psychological well-being in caregivers of individuals with mental illness: a cross-sectional study

**DOI:** 10.1101/2021.07.14.21260546

**Authors:** Edina Yi-Qin Tan, Vania T. Yip, Kim Lian Rolles-Abraham

**Affiliations:** Division of Social Sciences, Yale-NUS College, 16 College Avenue West, Singapore 138527; Better Life Psychological Medicine Clinic, 10 Sinaran Dr, #08-17/30, Singapore 307506

**Keywords:** Psychological well-being, Caregiving, Coping strategies, Benefit finding, Self-compassion

## Abstract

**Background:** Resilience factors that promote psychological well-being amongst caregivers of individuals with a mental illness are understudied. Coping strategies (problem-focused coping, emotion-focused coping and benefit-finding) have been found to influence the relation between stress and psychological outcomes, but their *relative* contribution to psychological well-being has not been tested. Self-compassion reduces caregiver burden in caregivers, yet no research has examined its contribution to psychological well-being nor the mechanisms via which it could do so. The aim of this study was to examine benefit-finding and self-compassion as resilience factors that could promote psychological well-being amongst caregivers of individuals with mental illnesses.

**Methods:** This cross-sectional study was conducted between June 2019 to October 2019 in Singapore. 107 informal caregivers of individuals with various diagnosed mental illnesses completed an online questionnaire. COPE, General Benefit Finding Scale (GBFS), Self-Compassion Scale -Short (SCS-SF), Psychological Well-Being Scale -Brief (PWBS-B), and Hospital Anxiety and Depression Scale (HADS) were used. Hierarchical multiple regression and mediation analyses were conducted.

**Results:** Benefit-finding was a more important predictor of psychological well-being compared to problem-focused and emotion-focused coping. Self-compassion was positively associated with psychological well-being amongst caregivers, and this is partially due to an increased use of problem-focused coping strategies.

**Conclusions:** Future interventions should cultivate caregivers’ benefit-finding and self-compassion and consider using psychological well-being as an additional outcome measure.

## Background

Approximately 1 in 5 individuals suffer from a mental illness in their lifetime [1]. The debilitating course of mental illness, coupled with increasing deinstitutionalization, means that informal caregivers such as family members typically take on the responsibility of caring for such individuals [2]. Due to the stressful nature of caregiving, prior studies on informal caregivers have predominantly focused on examining the risk factors associated with higher levels of caregiver psychopathology [2–4]. However, the field of psychology is increasingly interested in positive mental health, which involves having psychological well-being *in addition to* lacking symptoms of psychopathology [5]. Longitudinal studies have shown that psychological well-being may be able to buffer individuals against physical and mental illness over time [6,7]. Such studies, alongside studies demonstrating that caregiving involves positive aspects (e.g., improved family cohesion, receipt of gratitude and appreciation from care recipients) that might contribute to the psychological well-being of the caregiver [8–10], suggest that there is value in enhancing caregivers’ levels of psychological well-being. Hence, it is crucial to investigate the resilience factors that could sustain and boost psychological well-being amongst informal caregivers of individuals with mental illnesses.

## Coping Strategies

The most common theoretical model used for studying how caregivers cope with the stress of caregiving is Lazarus and Folkman’s stress-coping model [11,12]. According to the stress-coping model, how the caregiver appraises their stressors, as well as the coping strategies that they use to manage these stressors, plays a crucial role in determining the caregiver’s psychological well-being [13]. The two most well-studied coping strategies are emotion-focused and problem-focused coping strategies, both of which have been conceptualised as part of the stress-coping model [11] (Fig. 1). Emotion-focused coping refer to processes that individuals engage in to try to reduce the emotional distress generated by the stressor, while problem-focused coping refer to processes that individuals engage in to try to directly alter the situation positively [14]. Whether or not emotion-focused coping and problem-focused coping are adaptive ways of coping with the stressor tends to be highly dependent on the characteristics and specific context of the stressor. Problem-focused coping tends to be more adaptive in situations where something can be done, while emotion-focused coping tends to be more adaptive in situations that have to be accepted [15]. In cases when the stressor is favorably resolved using either of these two coping strategies, positive emotions such as happiness and pride are generated [11] (Fig. 1), with positive emotions in turn associated with psychological well-being [16].

**Figure 1.**
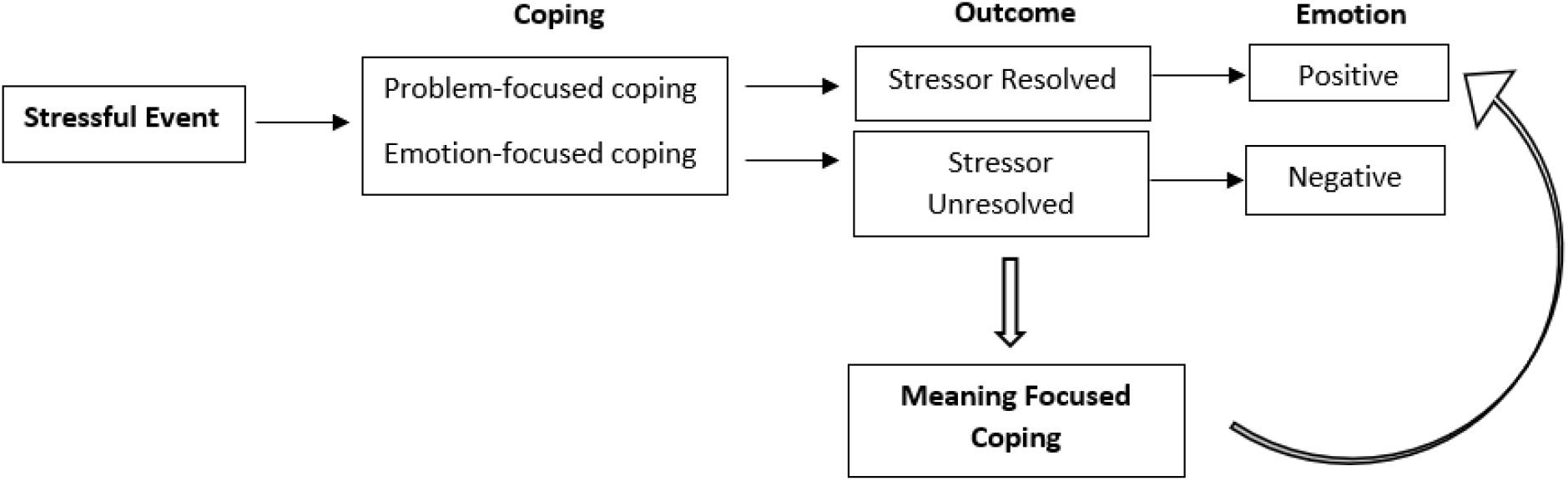
Simplified stress-coping model (Park & Folkman, 1997) *Note*. Meaning-focused coping has been hypothesized to generate positive emotions in situations where the stressor remains unresolved.

Considering mental illnesses are usually long-lasting with an unpredictable course [2], caring for an individual with a mental illness is typically a chronic stressor with no clear resolution [15]. This means that in the context of caregiving, traditionally well-studied adaptive coping strategies such as problem-focused coping and emotion-focused coping strategies may not be as effective in sustaining psychological well-being. This possibility is corroborated by inconsistent findings across studies which have examined the relations between problem-focused coping, emotion-focused coping, and caregiver psychological outcomes. Reliance on problem-focused coping has been associated with higher life satisfaction in prior caregiver studies [17], but may also lead caregivers to ignore the importance of taking time off from caregiving to focus on their own mental health [18]. Similarly, while some longitudinal studies have found that emotion-focused coping protects caregivers of individuals with mental illness from anxiety over time [19,20], others have found emotion-focused coping to be related to higher levels of depression in caregivers [21] — especially if the caregiver already had poor mental health and therefore decreased emotional regulation abilities [22].

In sum, problem-focused coping and emotion-focused coping may not adequately sustain psychological well-being in the context of caring for an individual with mental illness.

### Benefit-Finding

To explain the coping processes underlying irresolvable stressors, the stress-coping model was later revised to introduce a new category of coping, known as meaning-focused coping [23]. Meaning-focused coping is a form of appraisal-based coping whereby a person draws on his or her beliefs, values, and existential goals to motivate and sustain coping [23,24]. Coping by modifying one’s core beliefs, values, and goals allows an individual to generate positive emotions [15]. These positive emotions in turn influence the stress process by restoring psychological and social resources [16], thereby allowing individuals to sustain other types of adaptive coping, such as problem-focused coping and emotion-focused coping [23] (Fig.1).

The most common conceptualization of meaning-focused coping is benefit-finding [15], a construct typically assessed across several dimensions — growth in wisdom, patience and competence; greater appreciation for life, greater clarity about what matters, strengthened faith or spirituality, and improved quality of social relationships [15]. Benefit-finding has consistently been found to be positively associated with psychological well-being. A meta-analysis found a moderately strong positive relationship between benefit-finding and psychological well-being amongst the general population [25]. In the general caregiving literature, there is evidence that child caregivers (ages 12 to 16) of individuals with a range of mental, physical and developmental disabilities do experience benefit-finding, with benefit-finding being directly predictive of their psychological well-being [26]. Moreover, caregivers of individuals with Alzheimer’s disease who were assigned to a benefit-finding intervention not only reported lowered depressive symptoms relative to participants in control groups (i.e., no benefit-finding intervention), but also had increased levels of psychological well-being [27].

Hence, benefit-finding may be more important for sustaining psychological well-being in caregivers of individuals with mental illnesses compared to the more traditionally studied adaptive coping strategies; problem-focused coping and emotion-focused coping. As such, we hypothesize that benefit-finding will be positively associated with psychological well-being (H1), and benefit-finding will predict psychological well-being over and above emotion-focused coping and problem-focused coping strategies (H2).

### Self-Compassion

Compared to coping strategies, which are typically situation-dependent, a relatively more situation-independent resilience factor that could sustain psychological well-being in caregivers of individuals with mental illness is self-compassion. Self-compassion involves three components: (a) *self-kindness*, which involves being understanding towards oneself in the face of stress and failure, (b) *common humanity*, which involves viewing one’s suffering as part of a shared human experience wherein failures and imperfections are expected, and (c) *mindfulness*, which involves taking an open, accepting and nonjudgmental stance towards oneself and suffering [28].

Whether measured as a trait or a state, self-compassion has been found to relate positively to indicators of psychological well-being such as life satisfaction and subjective well-being, and relate negatively to anxiety and depression [29–31]. Importantly, self-compassion has been examined in the context of caregiving. Professional caregivers (i.e., nurses) who have high levels of self-compassion tend to provide more compassionate care and are more resistant to burnout compared to their less self-compassionate counterparts [32]. Informal caregivers follow a similar trend. Parents of children with Autism Spectrum Disorder who had higher levels of self-compassion experienced more life satisfaction and hope, and less stress and depressive symptoms [33]. Recently, [14] found that self-compassionate caregivers of individuals with dementia tended to experience lower levels of caregiver burden. Importantly, preliminary evidence suggests that self-compassion can be cultivated in caregivers. Caregivers of individuals with Alzheimer’s disease who participated in a yoga and compassion meditation program as part of an intervention study exhibited significant improvements in their levels of self-compassion post-intervention [34].

### Self-Compassion and Coping

A key mechanism via which self-compassion has been proposed to sustain psychological well-being during stressful times is by influencing persons’ use of coping strategies [14,31]. For instance, when faced with stressful circumstances, individuals higher in self-compassion have been found to experience less anxiety compared to those lower in self-compassion even after accounting for self-esteem [35]. Hence, self-compassion can be considered part of the stress coping process. The associations between self-compassion and emotion-focused coping, problem-focused coping, as well as benefit finding are outlined below.

#### Self-Compassion and Emotion-Focused Coping

A study that investigated this relationship found that self-compassionate students had a greater tendency to engage in emotion-focused coping strategies such as acceptance when encountering perceived academic failure [36], which led to greater positivity and resilience.

#### Self-Compassion and Problem-Focused Coping

Self-compassion is positively associated with variables that predict problem-focused coping, such as optimism and personal initiative [35]. Hence, caregivers who are high in self-compassion may be more likely to engage directly with the source of their stress rather than be a passive observer; enabling them to take on greater responsibility in attaining their personal goals [31]. Meaningful goal pursuit, in turn, has been found to relate positively to positive life outcomes, including psychological well-being [37,38].

#### Self-Compassion and Benefit-Finding

Self-compassion has been found to relate strongly to positive cognitive restructuring, which involves altering one’s view of a stressful situation to see it in a more positive manner [31]. Intervention studies have reported that positive cognitive restructuring helped participants view their stressors with greater self-directed compassion. Importantly, this increased self-compassion led to significant decreases in depression, anxiety and self-criticism amongst participants [35,39,40]. Given that benefit-finding involves a certain degree of positive cognitive restructuring [41], a possible pathway via which self-compassion may influence psychological well-being is by enhancing caregivers’ use of benefit-finding to cope with the stressors of caregiving.

Despite encouraging evidence, no research has examined whether self-compassion could enhance psychological well-being amongst caregivers of individuals with mental illness, nor the specific mechanisms via which self-compassion could influence psychological well-being. Hence, we propose two further hypotheses: Self-compassion will be positively associated with psychological well-being (H3) and the type of coping strategy used (problem-focused coping, emotion-focused coping, and benefit-finding) will mediate the relationship between self-compassion and psychological well-being (H4).

## Methods

### Ethics statement

Ethical approval for this study was obtained from the Institutional Review Board at the National University of Singapore.

### Study design and sample

The study sample consists of 107 caregivers who filled out an online survey between June 2019 and October 2019. Caregivers were recruited from two sites: a private clinic specialising in the treatment of eating disorders and a non-governmental organisation (NGO) dedicated to caregivers of individuals with mental illnesses^1^. For the private clinic, the recruitment process involved the distribution of flyers containing the link to the online survey to caregivers of persons attending the clinic. For the NGO, the recruitment process consisted of emails sent out to caregivers who had previously attended workshops held by the organisation. After providing informed consent, participants were told about the objectives of the study and subsequently completed an online survey. Upon study completion, participants were debriefed and received SGD $5 as reimbursement.

### Participant characteristics

All study participants were at least 21 years of age, and were informal caregivers of individuals with mental illnesses such as psychotic disorder, anxiety disorder, mood disorder, eating disorders, obsessive compulsive disorder, post-traumatic stress disorder, and unknown mental illnesses (wherein the caregiver did not know the specific name of the illness). Care recipient diagnoses were self-reported by the caregivers. To be eligible for the study, participants had to understand the English language, be the informal caretaker and/or relative of the person with mental illness and provide care and support to the individual with a mental illness. The informal caregiver was defined as an individual belonging to the care recipient’s informal support system who provides care for, is at least partly responsible for the individual, and spends a substantial amount of time on caregiving without financial reimbursement [2]. Most caregivers were aged between 51-60 (37.4%), with a preponderance of females (69.2%), Chinese (72.0%), university level education (65.4%), and employed (66.4%). A majority of the caregivers were mothers to the CR (37.4%), resided in the same household as their CRs (73.8%), and had other caregivers present in the same household (78.5%). The caregivers’ mean (SD) interaction hours (per week) with their CR was 33.1 (39.7). Most CRs were at least 20 years of age (82.7%), with a mean (SD) length of illness of 93.6 (135.1) months. The distribution of primary diagnoses amongst the CRs are as follows: mood disorder (35.5%), anxiety disorder (9.3%), psychotic disorder (23.4%), personality disorder (0.9%), eating disorder (22.4%), and other (8.4%). Participant characteristics are reported in Table 1.

**Table 1.**
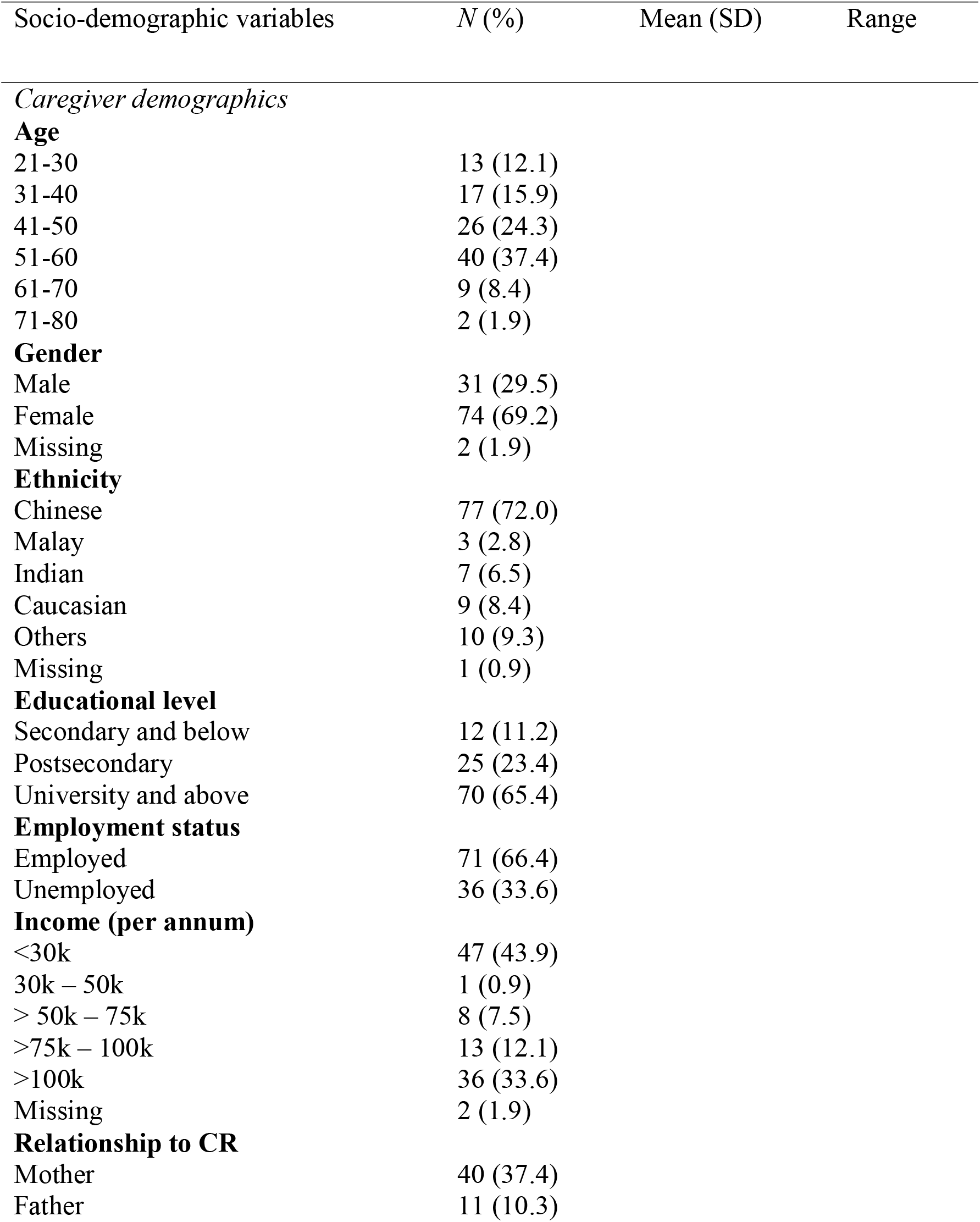
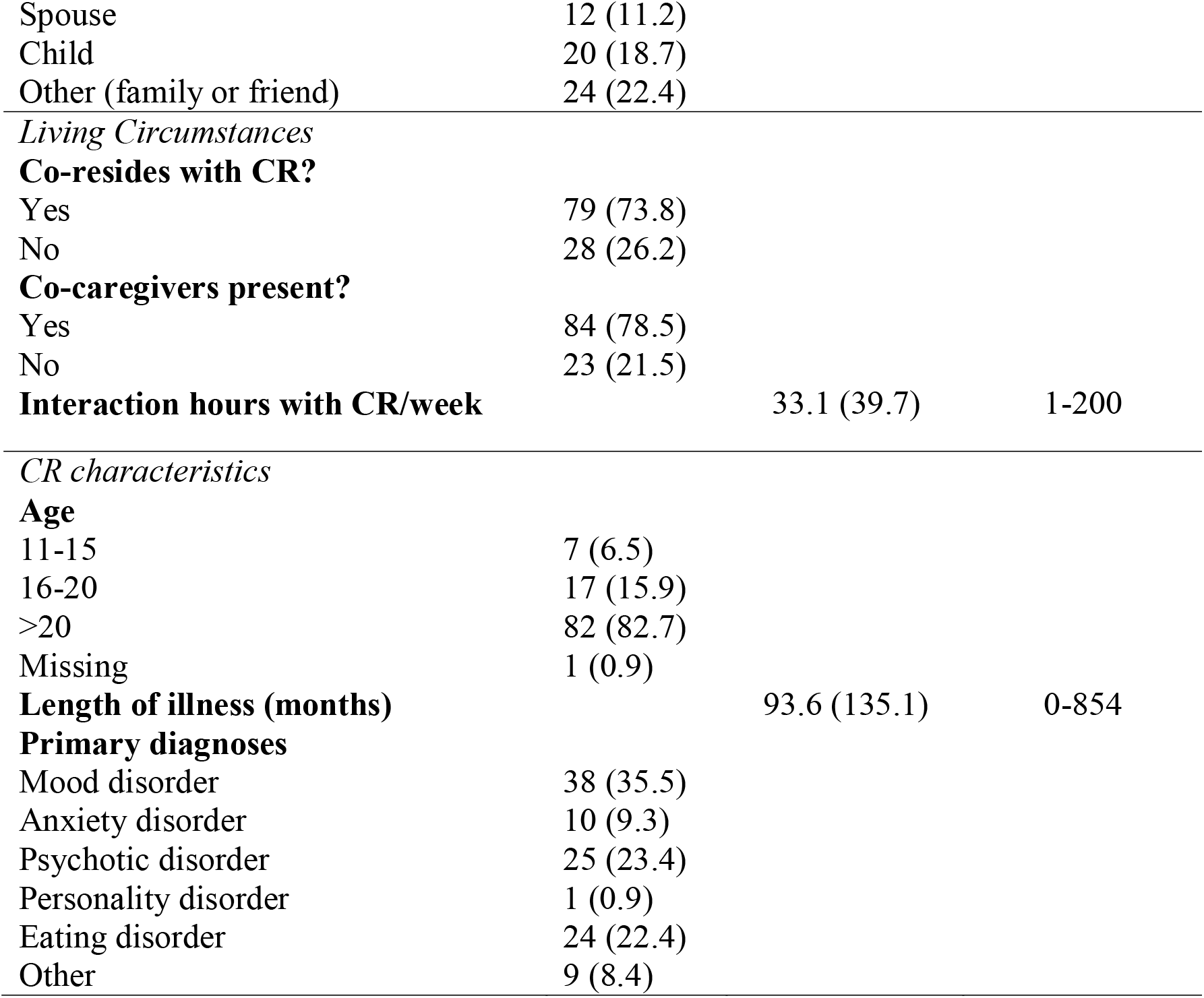
Participant characteristics.

### Measures

The questionnaire included the following instruments:

#### Socio-demographic Variables

Participants were asked for socio-demographic data such as age, gender, ethnicity, education level, employment status, income, living circumstances, age of care recipient (CR), CR diagnosis, length of CR illness, relationship to CR, and interaction hours with CR.

#### Problem-Focused and Emotion-Focused Coping Strategies

The 60-item Coping Orientations to Problems Experienced Inventory (COPE; [42]) were used to assess caregivers’ emotional-focused coping and problem-focused coping strategies. The COPE assesses 14 different coping strategies. These strategies can be subsumed under 3 categories: emotion-focused coping (acceptance, emotional support, humor, positive reframing, and religion), problem-focused coping (active coping, instrumental support, and planning), and dysfunctional coping (behavioral disengagement, denial, self-distraction, self-blame, substance use and emotional venting) [14,43]. The present study focuses on only emotion-focused and problem-focused coping, therefore only items belonging to these two categories were included. Participants were asked: ‘To what extent do the following statements describe how you cope with your present situation of caring for the individual with a mental illness?’ Example items include ‘I talk to someone to find out more about the situation’ (problem-focused coping) and ‘I accept that this has happened and that it can’t be changed’ (emotion-focused coping). Responses were recorded on a 4-point Likert-type scale where 1 = ‘I haven’t been doing this at all’ and 4 = ‘I’ve been doing this a lot.’ ‘Positive reframing’ items were not included in this study as positive reframing has been found to overlap statistically and conceptually with meaning-focused coping [17]. The rest of the items were summed to create an overall problem-focused coping and emotion-focused coping score. The COPE scale has been found to have good internal reliability, with an average Cronbach’s Alpha of *α* = .79 across all subscales [42]. In the present study, the internal reliability was α = .78 for emotion-focused coping and α = .89 for problem-focused coping.

#### Benefit-Finding

The 28-item General Benefit Finding Scale (GBFS; [44]) was used to assess benefit-finding. The scale incorporates six different factors and has been previously used to ascertain levels of benefit-finding in relation to general life stress (i.e., stress that is not personal trauma or illness-related). Participants were asked: ‘To what extent do the following statements describe your experience of caring for the individual with a mental illness?’ Example items include ‘Led me to be more accepting of things’ and ‘Helped me become a stronger person’. Responses were recorded on a 5-point Likert-type scale where 1 = ‘Strongly Disagree’ and 5 = ‘Strongly Agree’. All items were summed to create an overall benefit-finding score. The scale has been found to have good internal reliability, with Cronbach’s Alpha ranging from *α* =.76 to *α* = .86 across the six factors [45]. In the present study, the internal reliability was as follows: acceptance (*α* = .88), family bonds (*α* = .87), personal growth (*α* = .80), relationships (*α* = .87), empathy (*α* = .88) and reprioritisation (*α* = .79).

#### Self-Compassion

The 12-item Self-Compassion Scale - Short (SCS-SF; [46]) was used to assess self-compassion. Example items include ‘I try to see my failings as part of the human condition’ and ‘I’m disapproving and judgmental about my own flaws and inadequacies’. Responses were recorded on a 5-point Likert-type scale where 1 = ‘Almost Always’ and 5 = ‘Almost Never’. Negative items were reverse scored, and all items were summed to create an overall self-compassion score. The index has shown good internal consistency (α = .86) and has an almost perfect correlation with the full scale (r = .98) [14,46]. In the present study, the internal reliability was α = .75.

#### Psychological Well-Being

The 8-item Psychological Well-Being Scale -Brief (PWBS-B) was used to assess psychological well-being [47]. Example items include ‘I lead a purposeful and meaningful life’ and ‘I am engaged and interested in my daily activities.’ Responses were recorded on a 7-point Likert-type scale where 1 = ‘Strongly Disagree’ and 7 = ‘Strongly Agree’. All items were summed to create an overall psychological well-being score. The index has shown good internal consistency (α = .86) and showed strong correlations with the full scale (r = .80). In the present study, the internal reliability was α = .90.

#### Psychopathology

The 14-item Hospital Anxiety Depression Scale (HADS) was used to assess symptoms of psychopathology in the form of anxiety (7 items) and depressive states (7 items) [48]. Example items included ‘I still enjoy the things I used to enjoy’ (depression) and ‘I feel tense or wound up’ (anxiety). Participants rate the frequency of feelings over the past week on a scale from 0 = ‘Not at All’ to 3 = ‘Very Often’. In this study, the total HADS score was used, as there is evidence for a single dimension of general psychopathology in the HADS [6,49]. Higher scores indicate more symptoms of psychopathology (range 0–42). In the present study, the internal reliability was α = .90.

### Statistical Analysis

All statistical analyses were conducted using SPSS Version 20 (IBM SPSS Statistics). Missing values and outlier analysis were performed^2^. Residual and scatter plots indicate the assumptions of normality, linearity and homoscedasticity were all satisfied. Linear regression was performed to investigate if benefit-finding and self-compassion were significant predictors of psychological well-being. Hierarchical multiple regression analysis was performed to investigate if benefit finding predicted psychological well-being over and above problem-focused coping and emotion-focused coping. Mediation analyses were performed to explore potential mediators of the relationship between self-compassion and psychological well-being. For all analyses, statistical significance was set at *p* < 0.05 (two-tailed).

#### Power Analysis

Apriori power analysis was conducted using *G*Power 3*.*1* [50]. The power analysis indicated that a total sample of 103 participants will be needed to test medium effects (Cohen’s f^2^ = .15) with 80% power, using F-tests with alpha set at 0.05. Hence, the sample size of 107 in this study was deemed to have sufficient power to detect the effects of the resilience factors on psychological well-being.

## Results

This sample of caregivers exhibited low levels of psychopathology (i.e., depression and anxiety) overall, with a mean (SD) HADS score of 14.06 (7.83). Only one caregiver met the HADS criteria for clinical anxiety (HADS cut-off score of 21 or higher).

### Benefit Finding

A simple linear regression was carried out to investigate if benefit finding is a significant predictor of psychological well-being. Benefit finding was found to be a significant predictor of psychological well-being (F (8, 92) = 2.50, *p* = 0.017, β = .37, 95% CI [.10, .33]), with an R^2^ of .18 (Cohen’s f^2^ = .22) controlling for age, gender, ethnicity, education, income, CR diagnosis and relationship to CR. Previous caregiver studies have found these variables to covary with psychological well-being, and hence they were controlled for in the analyses (Trompetter et al., 2017; Lloyd et al, 2019). For this and all subsequent analyses conducted in this study, the same variables were controlled for.

A three-stage hierarchical multiple regression conducted with psychological well-being as the dependent variable revealed that problem-focused coping and emotion-focused coping (entered in step 2) accounted for 10.5% of the variation in psychological well-being, controlling for covariates (entered in step 1), Δ*F* (2, 94) = 4.00, *p* = 0.021. Benefit-finding accounted for an additional 8.9% of variance in psychological well-being when it was added to the model (step 3), with a significant change in R^2^ (Δ*F* (1, 93) = 10.25, *p* = 0.002, β *=* .34, 95% CI [.08, .32]). When all the independent variables were included in the regression model, only benefit-finding emerged as a significant predictor of psychological well-being. Hence, the most important predictor of psychological well-being was benefit-finding, which uniquely explained 8.9% of the variation in psychological well-being. Table 2 shows the results of the hierarchical multiple regression.

**Table 2.** Summary of hierarchical regression analysis

| Variable | $\beta$ | $t$ | R | $R^2$ | $\Delta R^2$ |
| --- | --- | --- | --- | --- | --- |
| **Step 1** |  |  | .17 | .03 | - |
| Age | 1.1 | 1.49 |  |  |  |
| Gender | -.10 | -.05 |  |  |  |
| Income | -.08 | -.16 |  |  |  |
| CR diagnosis | .30 | .59 |  |  |  |
| **Step 2** |  |  | .32 | .11 | .08 |
| Age | .89 | 1.24 |  |  |  |
| Gender | -.56 | -.29 |  |  |  |
| Income | -.01 | -.02 |  |  |  |
| CR diagnosis | .27 | .54 |  |  |  |
| Problem-focused coping | .36 | 2.59* |  |  |  |
| Emotion-focused coping | -.01 | -.06 |  |  |  |
| **Step 3** |  |  | .44 | .19 | .08 |
| Age | .57 | .85 |  |  |  |
| Gender | -.57 | -.31 |  |  |  |
| Income | .14 | .29 |  |  |  |
| CR diagnosis | .79 | 1.56 |  |  |  |
| Problem-focused coping | .24 | 1.93 |  |  |  |
| Emotion-focused coping | -.05 | -.42 |  |  |  |
| Benefit-finding | .20 | 3.20** |  |  |  |

Note. $N = 107$ , \* $p < .05$ , \*\* $p < .01$

### Self-Compassion

A simple linear regression was carried out to investigate if self-compassion is a significant predictor of psychological well-being. Self-compassion was found to be a significant predictor of psychological well-being (F (8, 94) = 2.31, *p* = 0.026, β = .33, 95% CI [.23, .95]), with an R^2^ of .16 (Cohen’s f^2^ = .20).

### Mediation Analyses

The fourth hypothesis predicted that coping strategies (emotion-focused coping, problem-focused coping, and benefit-finding) would be mediators of the relationship between self-compassion and psychological well-being. Problem-focused coping, emotion-focused coping, and benefit-finding were individually tested as the mediator across three separate models according to statistical and conceptual propositions for testing the presence of mediation [51]. Out of the three tested coping strategies, only problem-focused coping was significantly linked to self-compassion (B = .51, β = .36, *p* < 0.001, 95% CI [.25, .76]). Therefore, further analyses were conducted only for problem-focused coping. Problem-focused coping was also significantly associated with psychological well-being, controlling for self-compassion (B = .27, β = .22, *p* = 0.028, 95% CI [.03, .51]). For the above model including problem-focused coping as the mediator, the *z*-score was calculated to be 2.46 (95% CI [.08, .76]). Hence, problem-focused coping was a statistically significant partial mediator of the relationship between self-compassion and psychological well-being (Fig. 2). The analyses used to test the hypothesized models are presented in Table 3. Only problem-focused coping is presented in Table 3 as only problem-focused coping met the criteria for further mediation analyses.

**Table 3.** Testing for problem-focused coping as a mediator using multiple regression

| Steps in testing for mediation | B | SE B | 95% CI | $\beta$ | R<sup>2</sup> |
| --- | --- | --- | --- | --- | --- |
| *Problem-focused coping* |  |  |  |  |  |
| **Testing step 1 (path c)** | .55 | .16 | .23, .88 | .31** | .10 |
| Outcome: Psychological well-being |  |  |  |  |  |
| Predictor: Self-compassion |  |  |  |  |  |
| **Testing step 2 (path a)** | .51 | .13 | .25, .76 | .36** | .13 |
| Outcome: Problem-focused coping |  |  |  |  |  |
| Predictor: Self-compassion |  |  |  |  |  |
| **Testing step 3 (paths b and c')** | .27 | .12 | .03, .51 | .22* | .12 |
| Outcome: Psychological well-being | .42 | .17 | .08, .76 | .24* | .14 |
| Mediator: Problem-focused coping |  |  |  |  |  |
| Predictor: Self-compassion |  |  |  |  |  |

*Note.* CI Confidence Interval; \* $p < .05$ , \*\* $p < .01$ **Discussion**

**Figure 2.**
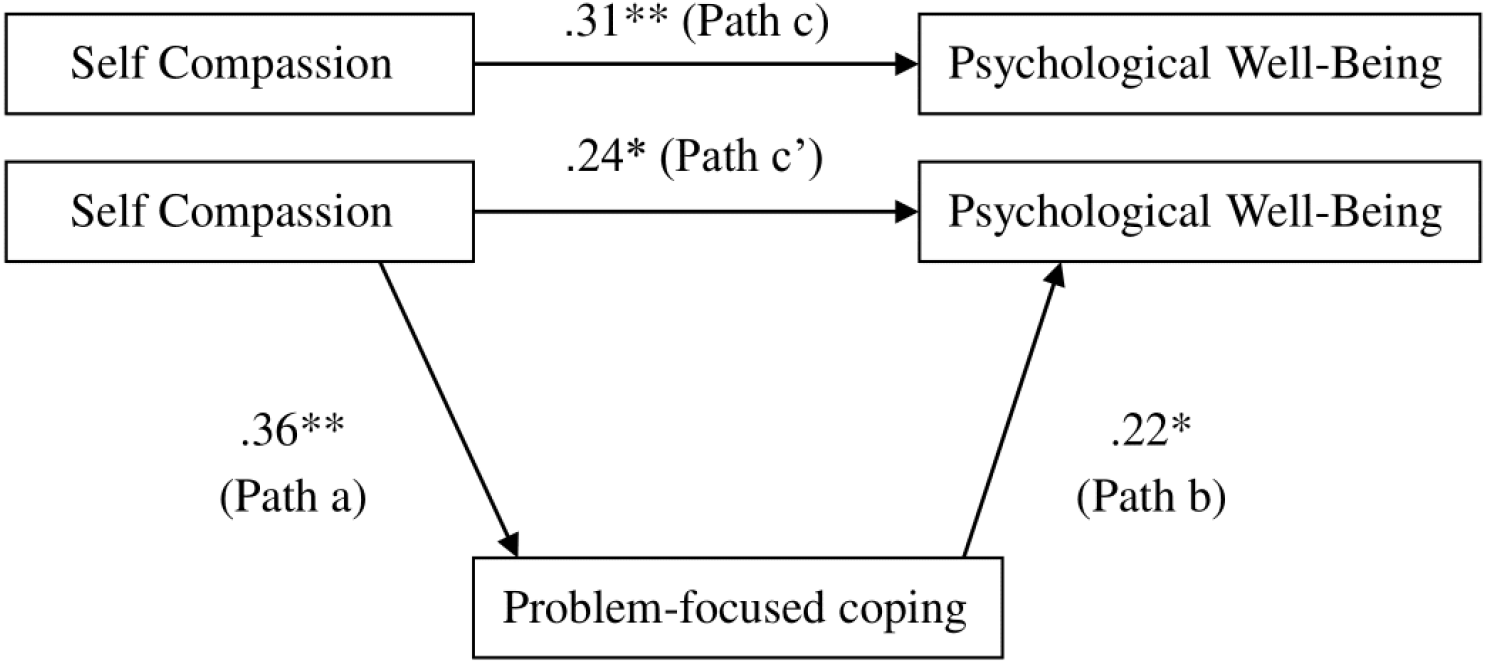
Testing for problem-focused coping as a mediator *Note*. Standardized regression coefficients for the relationship between self-compassion and psychological well-being as mediated by problem-focused coping. \**p* < .05, \*\**p* < .01

## Discussion

The present study aimed to examine benefit-finding and self-compassion as resilience factors that could promote psychological well-being in caregivers of individuals with mental illnesses. As predicted by the first hypothesis, benefit-finding was significantly associated with caregiver psychological well-being. Caregivers who reported high levels of benefit-finding experienced higher levels of psychological well-being than those lower in benefit-finding. Moreover, in line with the second hypothesis, benefit-finding predicted psychological well-being over and above the traditionally studied adaptive coping strategies, problem-focused coping and emotion-focused coping. These results demonstrate that for caregivers of individuals with mental illness, benefit-finding is a more important contributor to psychological well-being than emotion-focused coping or problem-focused coping. The results of the present study are consistent with, and add to, [23] revised stress coping model. Specifically, these results provide the first empirical evidence for the previously untested theory that meaning-focused coping strategies, such as benefit-finding, may play a relatively more crucial role in sustaining psychological well-being under circumstances where the stressor is chronic and unresolvable— such as caring for individuals with long-term mental illnesses.

The third hypothesis was also upheld, with self-compassion being positively associated with caregiver psychological well-being. The results are consistent with previous research, which found that self-compassion was negatively associated with *caregiver burden* among caregivers of individuals with dementia [14]. However, this study is the first to show that self-compassion and psychological well-being are positively associated in caregivers of individuals with a mental illness.

The fourth hypothesis, which predicted that the relationship between self-compassion and psychological well-being would be mediated by the use of adaptive coping strategies (emotion-focused coping, problem-focused coping and benefit-finding), was partly upheld. This study found that problem-focused coping partially mediates the relationship between self-compassion and psychological well-being. These results show that caregivers high in self-compassion have a greater tendency to utilise problem-focused coping strategies, which in turn at least partly leads to increased psychological well-being. This is consistent with previous studies, which found that self-compassionate caregivers are more likely to use adaptive coping strategies [14,52,53].

In this study, emotion-focused coping was not a significant mediator of the relationship between self-compassion and psychological well-being. This finding is consistent with and extends a previous study which found that emotion-focused coping does not appear to mediate the relationship between self-compassion and caregiver burden [14]. Emotion-focused coping strategies have been found to function differently compared to problem-focused coping and meaning-focused coping in the stress-coping model (Fig. 1). Specifically, emotion-focused coping tends to have complex, unpredictable interactions with caregiver outcomes and other caregiving factors. For instance, while some research has found that adopting emotion-focused coping strategies protect caregivers against anxiety [19], others have found no such relationship [14,54]. Hence, further research is warranted to elucidate the role of emotion-focused coping in relation to self-compassion and psychological well-being.

Surprisingly, benefit-finding was also not a significant mediator of the relationship between self-compassion and psychological well-being. This finding is unexpected, considering that (a) meaning-focused coping has been indirectly linked to self-compassion [31,41] and (b) greater reliance on meaning-focused coping has been associated with lower distress in prior caregiver studies [17]. However, it should be noted that the measures used in this study differ from measures used in previous caregivers studies. Prior studies tended to focus on caregiver distress, rather than psychological well-being as the outcome. However, the absence of caregiver distress is conceptually and functionally distinct from the presence of psychological well-being [55]. Hence, different mechanisms may underlie the relationship between benefit-finding, self-compassion and the outcome of psychological well-being, as compared to the outcome of caregiver distress. For instance, adopting benefit-finding coping strategies may lower caregivers’ levels of psychological distress without necessarily increasing their levels of well-being [55]. Future research should use consistent measures to better understand the mechanisms relating self-compassion, benefit-finding, and psychological well-being.

### Clinical Implications

The finding that benefit-finding and self-compassion are positively associated with the psychological well-being of caregivers with mental illnesses highlights an important gap in current clinical care for caregivers. Beyond assessing symptoms of psychopathology amongst caregivers, this study shows that psychological well-being is also an effective measure of caregivers’ quality of life and should be incorporated into current clinical assessments of caregivers to increase their ability to identify gaps in support. The significant relationships between the resilience factors, benefit-finding and self-compassion, to psychological well-being also highlight the importance of assessing these resilience factors. Specifically, low levels of benefit-finding and/or self-compassion could serve as reliable indicators of caregivers who are currently experiencing, or at risk of experiencing, low levels of psychological well-being.

Future intervention designs for caregivers could also benefit from the findings of this study. Although interventions promoting self-compassion and benefit-finding are gaining popularity and increasingly recognized as effective and accessible ways of supporting at-risk individuals [40,56], they tend to be designed with the goal of alleviating psychological distress rather than the goal of boosting psychological well-being. While alleviating psychological distress is an undoubtedly crucial goal, incorporating the goal of enhancing psychological well-being could go a long way in changing the mindsets of not only the caregivers, but also the clinicians and researchers involved. Specifically, interventions could incorporate content that encompass the multifaceted components of psychological well-being (e.g., positive social relationships, environmental mastery) and be held over a longer period of time, even after psychological distress has been measurably reduced. Moreover, scales that measure psychological well-being can be utilized alongside scales that measure psychological distress in order to add a further impact measure for the efficacy of current interventions.

The findings of this study should be viewed in light of its limitations. Firstly, analyses by subgroups of caregivers (e.g. by CR diagnoses) could not be conducted due to an insufficient sample size across sub-groups. Future studies can focus on at-risk subgroups of caregivers that have been identified as experiencing lower benefit-finding, self-compassion and/or psychological well-being. Secondly, the cross-sectional design of this study limits any ascertainment of causality. Future longitudinal studies could be run to provide stronger evidence for the causal relationship between benefit-finding, self-compassion and psychological well-being.

## Conclusions

The aim of the present study was to explore the role of benefit-finding and self-compassion in promoting psychological well-being amongst caregivers of individuals with mental illnesses. The results of the study provide first empirical evidence that benefit-finding and self-compassion are crucial coping strategies that can promote caregivers’ psychological well-being. However, more research is needed to explore whether and how the findings from this general study could apply to specific sub-groups of caregivers, as well as to explore the causal relations between benefit-finding, self-compassion, psychological well-being and psychopathology. Regardless, the present findings provide promising empirical evidence which can be used to develop interventions for caregivers that centre not only on reducing psychological distress, but also promoting psychological well-being. Such interventions could equip caregivers with the mental resilience necessary to shoulder their hefty caregiving responsibilities, while still living a flourishing life.

## Data Availability

The datasets used and/or analysed during the current study are available from the corresponding author upon reasonable request.

## Declarations

### Ethics approval and consent to participate

Ethical approval was obtained from the Institutional Review Board at the National University of Singapore. All study participants provided informed consent prior to taking part in the study.

### Consent for publication

Not applicable

### Authors’ contributions

EYQT, VTY and KLRA conceptualized the study. EYQT analyzed the data and wrote the manuscript. VTY was a major contributor in writing the manuscript. All authors read and approved the final manuscript.

## Acknowledgements

Not applicable

## Footnotes

1 These two samples did not differ significantly on the constructs of interest (i.e., coping strategies, self-compassion, psychological well-being, psychopathology), and were therefore combined. All analyses that follow are based on this combined set of caregivers.

2 On average, 1.87% of data was missing due to inconsistent reporting by participants. In total, 26% of participants (n = 28) had one or more missing data points. Outlier analysis revealed potential outliers in PWBS-B (n = 2), GBFS (n = 7) and SCS-SF (n = 7). As removing these outliers did not change the results of any analyses performed, only results with outliers included will be shown and discussed in the remainder of this article.

